# The Link between Poverty and COVID-19 Case and Mortality Rates in Germany

**DOI:** 10.1101/2020.08.09.20171207

**Authors:** Felix Ettensperger

## Abstract

The effects of poverty on the case and mortality rates of Covid-19 has emerged as a controversial but understudied topic. In previous studies and reports from the UK and US evidence emerged that poverty related indicators had a significant statistical effect on case and mortality rates on district level. For Germany, it has largely been assumed that poverty is an equally relevant factor influencing the transmission rates of the outbreak mostly due to anecdotal evidence from local outbreaks in meat processing plants and reported incidents in poorer city districts. This paper addresses the lack of statistical evidence and investigates thoroughly the link between poverty related indicators and case and mortality rates of the outbreak using multivariate, multilevel regression while also considering the urban-rural divide of the country. As proxies for poverty the unemployment rate, the per capita presence of general practitioners (physicians), per capita GDP, and the rate of employees with no professional job training is evaluated in relation to the accumulated case and mortality numbers on district level taken from RKI data of June and July 2020. Interestingly, the study finds no evidence for a poverty-related effect on mortality for German districts. Furthermore, only employment in low qualification jobs approximated by the job training variable consistently affected case numbers in urban districts in the expected direction.

## Research Focus

The link between poverty and Covid-19 infection and mortality rates has been intensively discussed in international articles and media in recent weeks. A variety of scientific papers has been released with evidence for poverty related effects on the infection risk and mortality rates in different countries. However, a systematic analysis for the effect of poverty during the Covid-19 pandemic in Germany is still lacking. This is a research gap addresses by this paper.

Poverty is suspected to be a risk factor for increased infection rates during a pandemic like the current Covid-19 outbreak for a variety of reasons. Accommodations in poorer urban zones are usually on average smaller and population density is much higher in poor urban neighborhoods compared to affluent ones (McFarlane 2020; Cooke 1999). This limits the possibility for social distancing. Furthermore, workers in precarious types of employment are less likely to continuously work within a safe distance to co-workers and clients, less able to take payed sick leave and are thus suspected to be more prone to infection risks in many industries and occupations (ILO 2020; Finch and Hernández Finch 2020; Burgard and Lin 2004). A series of spreading events in precarious employed workers in the meat and agriculture industry in Germany, the US and other countries have highlighted this risk factor. Jobs in low education employment, mostly in manual repetitive labor, also in general cannot be in done in any sensible way in a home office setup to protect employees. Most workers in these sectors have a higher dependence on public transport (Rachele et al. 2015) to reach their work site and allegedly sub-standard access to Personal protective equipment (PPE). The third set of reasons why poor communities and groups might be more prone to infection risk is the lack of trust in official information about infection risks provided by experts. Higher Income is positively related to trust in media (Kalogeropoulos et al. 2019) and poorer social groups might be more prone to misinformation provided by unqualified sources via social media and disinformation agents.

Poverty is also, for a number of reasons, assumed to be indirectly linked to higher mortality rates and more severe health outcomes for the infected. Especially for the US uneven health care coverage and unaffordable access to health services which lead to a higher prevalence of chronic illness among poorer citizens has been discussed as a potential reason why poverty is linked to a higher risk of death caused by Covid-19 in recent studies (Blumenthal et al. 2020). But, also in the UK, people living in deprived areas experienced mortality rates more than double the rate of affluent areas (Caul 2020). Studies also found the likelihood of intergenerational households to be more likely in less affluent districts in the UK, a fact that might limit the possibility of social distancing and cause more severe infection rates among vulnerable demographic groups and more frequent negative health outcomes in general (Kenway and Holden 2020). It is also likely that the higher prevalence of obesity among poorer citizens is negatively affecting survival rates of patients. This is highly plausible due to the fact, that obesity has a very significant effect on health complications related to Covid-19 infections in clinical studies (Petrilli et al. 2020) and is in a multitude of developed countries clearly connected to higher poverty and lower average education as demonstrated in several studies (Levine 2011, Mensink et al. 2013).

In US Studies there is also an ongoing discussion and observations of an independent statistical effect (after controlling for poverty) for higher case and mortality rates among disadvantaged ethnic minorities (Gaglioti et al. 2020). This connection can simply not be examined in Germany due to the lack of available data for ethnicity on the district level.

The focus in this study is unidirectional. The pandemic is likely to cause more poverty due to the socio-economic impact of lockdowns, fear, and strained medical systems in the future, as well as in the short term. Certainly, looking at this reverse effect is a valid point of research, but this paper will exclusively focus on the effect of poverty and poverty related variables enabling and causing more infection cases and deaths during the pandemic, not vice-versa.

### Previous research

Most evidence for the connection of infection cases, mortality and poverty indicators is coming from the United Kingdom and the US. In one study investigating the phenomenon on district level the authors Finch and Hernández Finch (2020) found for the collected US Data a solid connection between poverty and infection cases as well as for poverty and the mortality data. According to their research investigating the time from February 15 to April 1 in 2,853 of the 3,007 counties the less affluent, disadvantaged districts suffered disproportionally higher case and mortality numbers.

They applied data from the Poverty Solutions Initiative (PSI) based on 5 indicators of poverty including estimate of social mobility, life expectancy, percent of residents living below the poverty line, percent of residents living in deep poverty, and the percent of low birth weights (Finch and Hernández Finch 2020: 3, Robles et al. 2020).

Especially in the beginning the effect they discovered was very pronounced. Interestingly, the trend reversed for their last measurement date, indicating that at a later stage more affluent communities began to see higher case numbers as well. The authors even argued that the late response to the pandemic was connected to the fact, that initially affluent districts remained relatively unaffected in the beginning. For mortality the authors discovered a link between poverty and mortality especially for the last measurement date on April 1st 2020.

In the statistical bulletin of the Office for National Statistics created on June 12 by Sarah Caul (2020) the connection between poverty and age-adjusted mortality rates is highlighted for the UK. Caul also separates the data in urban and rural districts based on the Urban and Rural Area Definitions of the UK Department of Environment, Food & Rural Affairs. She finds urban major conurbation (foremost the metropolitan area of London) to have a significantly increased age-standardized mortality rate compared to any other type of district. Also, by connecting the mortality data with the Welsh and English Index of Multiple Deprivation, she finds clear evidence for a connection between poverty and Covid-19 mortality in England and Wales. Mortality was more than double for poorer districts compared to affluent ones. The data used in her research was covering the timeframe between 1 March 2020 and 31 May 2020. Both studies see a relatively clear connection between poverty and infection risks as well as more serious health outcomes for the infected poor.

## Research Setup and Data

The study design to test these assumptions for Germany is divided into two parts: The first part is concerned with 400 of the 412 different regions in Germany covering all rural county districts (Landkreise, Kreise) and all urban districts (kreisfreie Städte and Stadtkreise) excluding only the 12 capitol districts (Bezirke) of Berlin due to incompatible socio-economic data for these specific districts. Socio-economic data including unemployment, district GDP, rate of general physicians and the rate of employment without professional qualification is collected from official data sources including the federal employment agency (Bundesagentur für Arbeit) and from datasets developed and collected by the Ministry of Agriculture and Rural Development. A descriptive summary of the poverty indicators is given in Table 1.

**Table 1.**
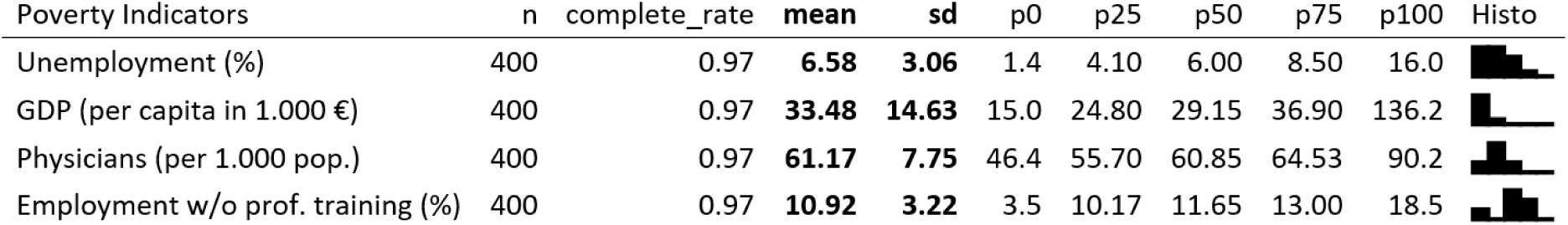

| Poverty Indicators | n | complete_rate | mean | sd | p0 | p25 | p50 | p75 | p100 | Histo |
| --- | --- | --- | --- | --- | --- | --- | --- | --- | --- | --- |
| Unemployment (%) | 400 | 0.97 | **6.58** | **3.06** | 1.4 | 4.10 | 6.00 | 8.50 | 16.0 |  |
| GDP (per capita in 1.000 €) | 400 | 0.97 | **33.48** | **14.63** | 15.0 | 24.80 | 29.15 | 36.90 | 136.2 |  |
| Physicians (per 1.000 pop.) | 400 | 0.97 | **61.17** | **7.75** | 46.4 | 55.70 | 60.85 | 64.53 | 90.2 |  |
| Employment w/o prof. training (%) | 400 | 0.97 | **10.92** | **3.22** | 3.5 | 10.17 | 11.65 | 13.00 | 18.5 |  |

A classification of districts based on rurality by the Thünen-Institute, a research institution and part of the federal department of agriculture is included to divide the data into different types of districts based on their degree of rurality and affluency. The rurality data is separating the districts into the following 5 categories: urban zone, rural zone (economically affluent), rural zone (economically marginalized), semi-rural (economically affluent) and semi-rural (economically marginalized).

Berlin is analyzed separately in detail in the second part of the analysis by district level (Bezirksebene) and with the help of detailed socio-economic data from the report “Monitoring Soziale Stadtentwicklung 2019” which is continuously developed and published annually since 1998 by the city planning office (Senatsverwaltung für Stadtentwicklung und Wohnen). The report contains detailed and very recent information about unemployment and social transfers on district and on subdistrict level for Berlin.

Covid-19 related data, including the variables cases, deaths, cases per 100k inhabitants, cases per 100k inhabitants (in the last 7 days) and death rate are directly taken from the Robert-Koch-Institute’s ArgCis Data repository and have been accessed during different dates to check for changes in distributional patterns during different moments of the pandemic and to update the data continuously.

## Results from German Districts

First, the connection between registered case numbers and poverty indicators is thoroughly investigated. To provide a general overview over the available data a linear multivariate regression model with the dependent variable accumulated cases per 100k inhabitants is conducted including all 400 districts (excluding Berlin). The results are shown in table 2. As independent variables to explain cases are included: unemployment, GDP, number of general physicians and percentage of employment without professional qualification (model 1).

**Table 2.**

| Socioeconomic effects on the case rate |  |  |  |  |  |  |
| --- | --- | --- | --- | --- | --- | --- |
|  | **Model 1** | **Model 2** | **Model 3** | **Model 4** | **Model 5** | **Model 6** |
|  | **All Districts** | **Urban Districts** | **Rural Districts** | **All Districts** | **Urban Districts** | **Rural Districts** |
|  | **June 14, 2020** | **June 14, 2020** | **June 14, 2020** | **July 7, 2020** | **July 7, 2020** | **July 7, 2020** |
| (Intercept) | 22.226<br>(75.569) | -26.649<br>(96.832) | 77.511<br>(95.648) | 31.899<br>(77.370) | -25.623<br>(101.242) | 93.659<br>(97.582) |
| unemployment | -20.953***<br>(2.759) | -10.854**<br>(3.232) | -27.610***<br>(4.137) | -20.237***<br>(2.824) | -9.109**<br>(3.379) | -27.466***<br>(4.221) |
| GDP | 0.776<br>(0.519) | 0.893<br>(0.460) | 1.343<br>(1.047) | 1.006<br>(0.531) | 1.049*<br>(0.481) | 1.801<br>(1.068) |
| physicians | 3.988***<br>(0.976) | 2.995*<br>(1.180) | 4.347***<br>(1.238) | 3.611***<br>(1.000) | 2.666*<br>(1.234) | 3.913**<br>(1.263) |
| no_prof_training | 6.475*<br>(2.684) | 8.972**<br>(3.301) | 1.303<br>(3.897) | 7.440**<br>(2.748) | 9.982**<br>(3.451) | 1.687<br>(3.976) |
| R<sup>2</sup> | 0.249 | 0.312 | 0.259 | 0.242 | 0.277 | 0.258 |
| Adj. R<sup>2</sup> | 0.242 | 0.282 | 0.250 | 0.234 | 0.244 | 0.248 |
| Num. obs. | 400 | 95 | 305 | 400 | 95 | 305 |

\*\*\* p < 0.001; \*\* p < 0.01; \* p < 0.05

In model 2 the same variables are tested only for the 96 urban districts. In model 3 the 304 rural districts are included. Models 1-3 contain data from mid-June 2020, Model 4-6 contains updated data from early July 2020. Model 4 is again for all districts, while Model 5 and 6 again separate into urban and rural districts with the updated July data. Table 3 contains VIF measures as control for multicollinearity in the models. The VIF values remain in a range which is generally considered unproblematic for multivariate models.

**Table 3.** Control for multicollinearity - VIF Values

|  | unemployment | GDP per capita | physicians | % with no prof. job training |
| --- | --- | --- | --- | --- |
| Model 1, June 14 - VIF | 1.335 | 1.116 | 1.335 | 1.116 |
| Model 2, urban - VIF | 1.079 | 1.109 | 1.079 | 1.109 |
| Model 3, rural - VIF | 1.07 | 1.859 | 1.07 | 1.859 |
| Model 4, July 7 - VIF | 1.398 | 1.217 | 1.398 | 1.217 |
| Model 5, urban - VIF | 1.192 | 1.114 | 1.192 | 1.114 |
| Model 6, rural - VIF | 1.299 | 2.006 | 1.299 | 2.006 |

The estimates are visualized in figure 1 for the first date and in figure 2 for the second measurement date.

**Figure 1.**
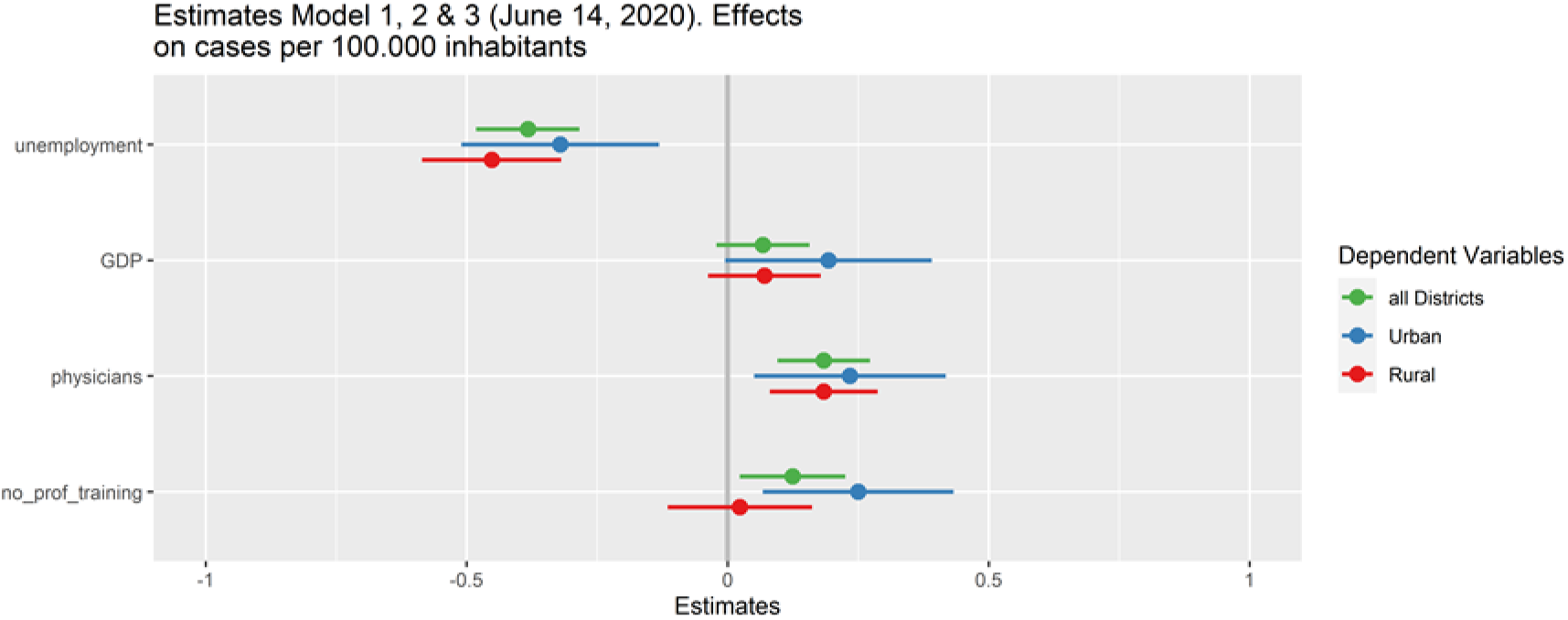

**Figure 2.**
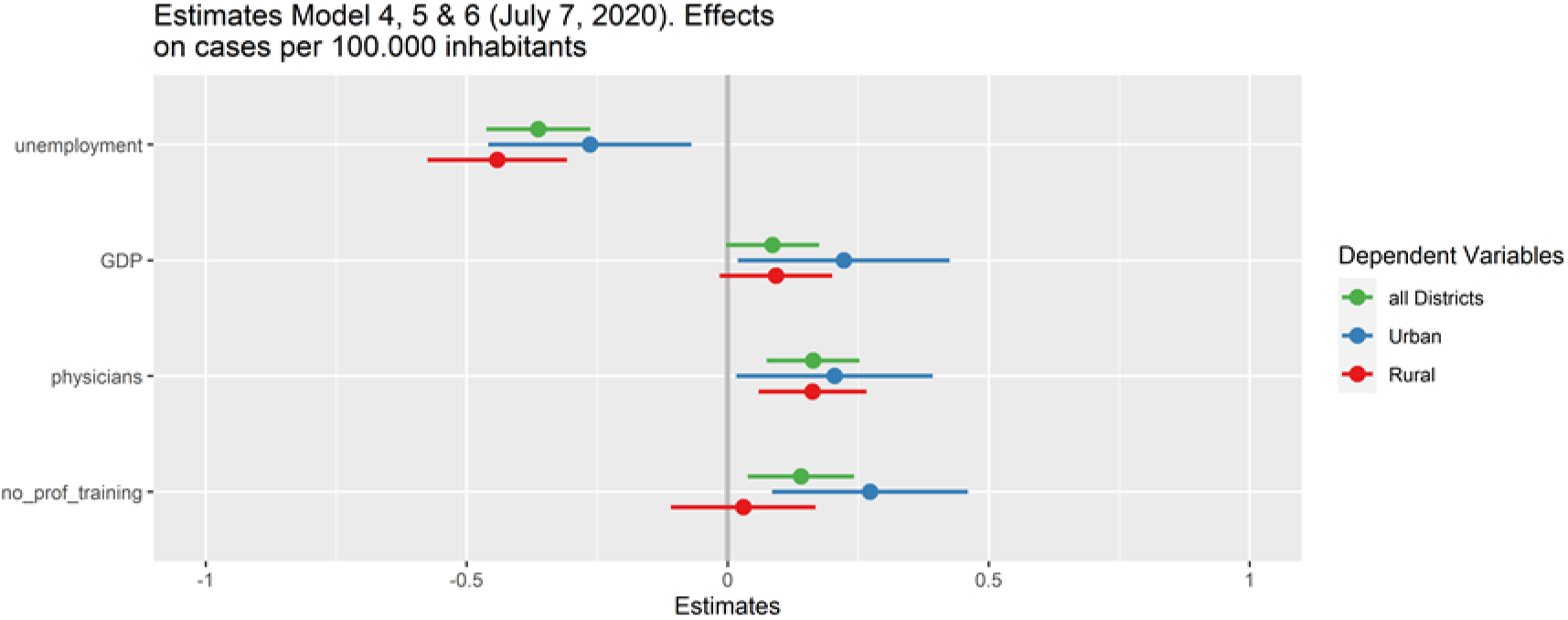

As opposed to what we might expect based on studies from the USA and UK, unemployment as a general proxy for poverty is not associated with increasing case numbers in any of these models, at least not on the investigated district level. In Germany, for all districts, and for urban and rural districts separately, higher unemployment figures consistently relate to lower Covid-19 cases. This connection remains true for different measurement dates.

At first glance contradictory, these results make sense if we check for the geographical distribution of cases: the worst affected districts in the beginning of the outbreak in Germany were mostly in the richer southern parts of Germany including many places in Bavaria and Baden-Württemberg (figure 3) with very low unemployment figures and a high density of general practitioners. Many poor and rural parts in eastern Germany with an opposite unemployment and physician constellation, on the other side, maintained low infection numbers throughout the first phase of the outbreak.

**Figure 3:**
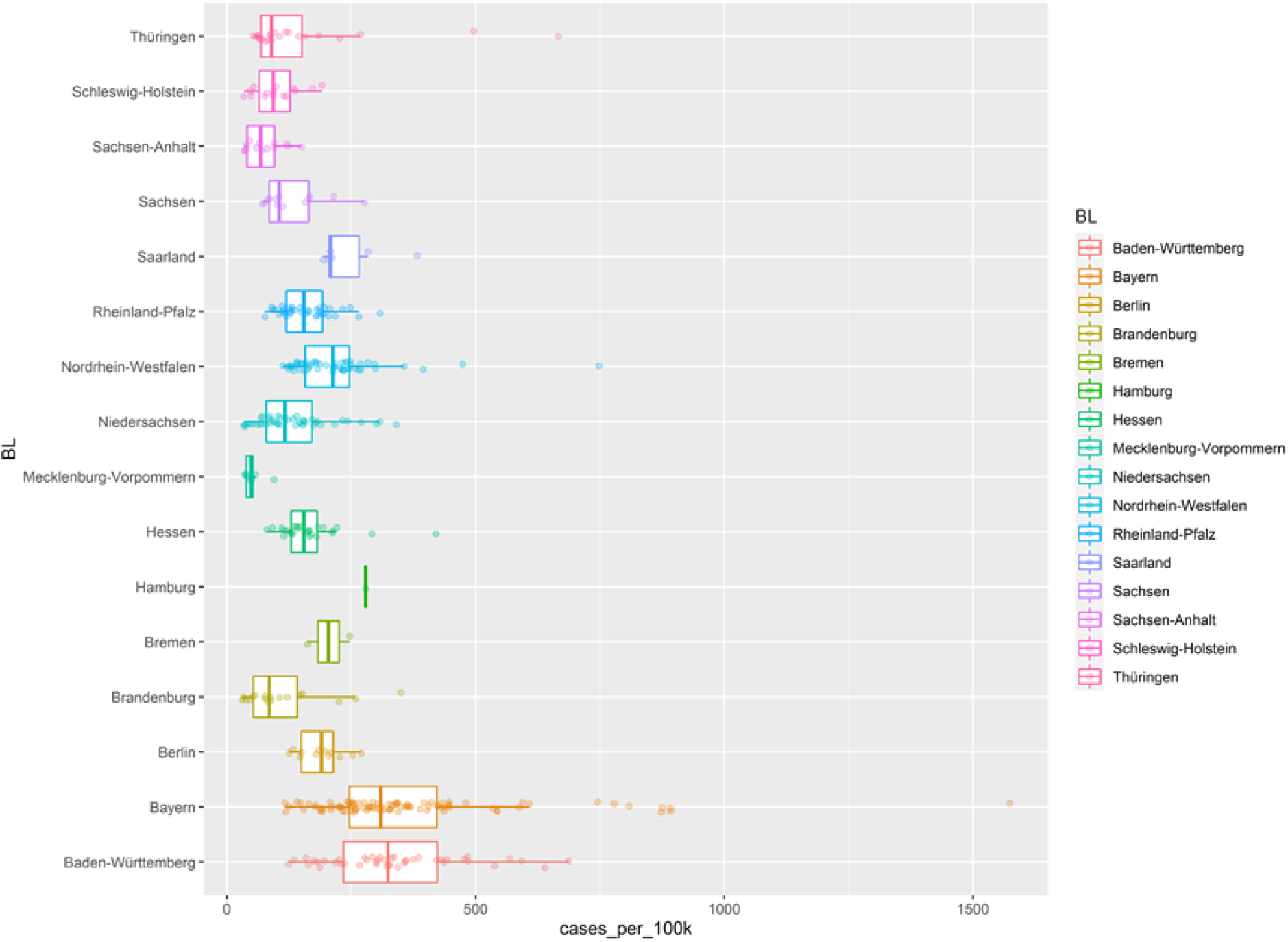
Cases per 100.000 inhabitants (divided by Bundesland), Date: June 14, 2020

The differences between north-south and east-west divide is investigated by using regional dummy variables which are included in separate regression model shown in table 4.

**Table 4.**

|  | Model 7<br>All Districts<br>June 14, 2020 | Model 8<br>All Districts<br>June 14, 2020 | Model 9<br>All Districts<br>July 7, 2020 | Model 10<br>All Districts<br>July 07, 2020 |
| --- | --- | --- | --- | --- |
| (Intercept) | 9.054<br>(76.319) | 60.950<br>(70.298) | 18.565<br>(78.141) | 69.427<br>(72.585) |
| unemployment | -22.278***<br>(2.970) | -5.890<br>(3.173) | -21.579***<br>(3.041) | -5.640<br>(3.276) |
| GDP | 0.001<br>(0.001) | 0.000<br>(0.000) | 0.001*<br>(0.001) | 0.000<br>(0.001) |
| physicians | 4.090***<br>(0.979) | 1.312<br>(0.965) | 3.714***<br>(1.003) | 1.018<br>(0.997) |
| no_prof_training | 11.236*<br>(4.787) | 6.015*<br>(2.491) | 12.260*<br>(4.901) | 6.994**<br>(2.572) |
| West | -51.452<br>(42.849) |  | -52.086<br>(43.872) |  |
| South |  | 160.082***<br>(19.916) |  | 155.133***<br>(20.564) |
| R<sup>2</sup> | 0.252 | 0.355 | 0.244 | 0.337 |
| Adj. R<sup>2</sup> | 0.243 | 0.347 | 0.235 | 0.329 |
| Num. obs. | 400 | 400 | 400 | 400 |

\*\*\*p < 0.001; \*\*p < 0.01; \*p < 0.05. West = including all Western states from before re-unification, South = including the 2 southern most states of Germany: Baden-Württemberg and Bayern Statistical models

As evaluation of the geographical effect it can be summarized that the north-south divide has a better explanative power than the east-west divide. Including a north-south dummy variable, the R-squared values of the model increase considerably and the variable’s effect on the model is highly significant, obscuring even the impact of the other indicators by rendering them insignificant. The east-west difference however, while being significant in a t-test, remains insignificant in the regression model, indicating, that the east-west difference, often associated to the former divide of the country, was far less important to explain distinctive transmission patterns in covid-19 cases.

Returning to the main models, it is interesting to observe that the number of low qualified employees (expressed by the variable of employees without professional qualification) is significant for model 2 and model 5, which are only including the urban districts. Apparently, for urban zones, the number of low qualified employees was connected to more covid-19 cases. Simultaneously, there was still a negative effect for unemployment on case numbers in the very same districts. This indicates that especially employment in precarious jobs is a risk factor for higher case levels.

For rural zones, on the other hand, this effect of the “percentage of jobs without professional qualification”-variable vanishes. This indicates that employment in low-qualification work was mainly in an urban context relevant for increasing the propensity of infections in Germany, not in rural districts. ^1^

In a multi-level-regression model, separating between urban and four groups of rural districts based on the Thünen-type, we clearly see that less dynamic regions with lower economic success did experience significantly less infection numbers. This is true both for rural and semi-rural zones. The distribution of cases between Thünen-types is shown in figure 4.

**Figure 4.**
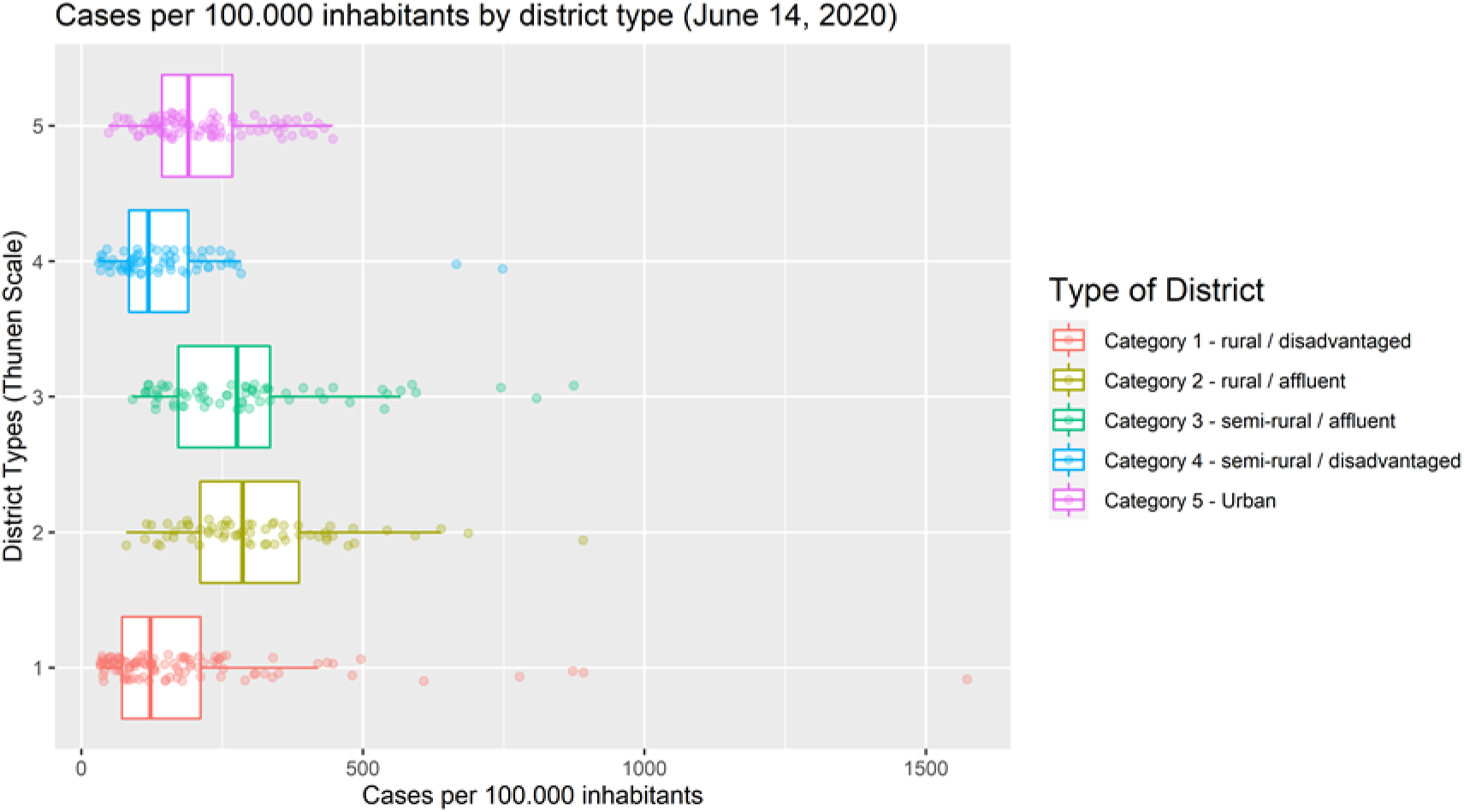

**Figure 5:**
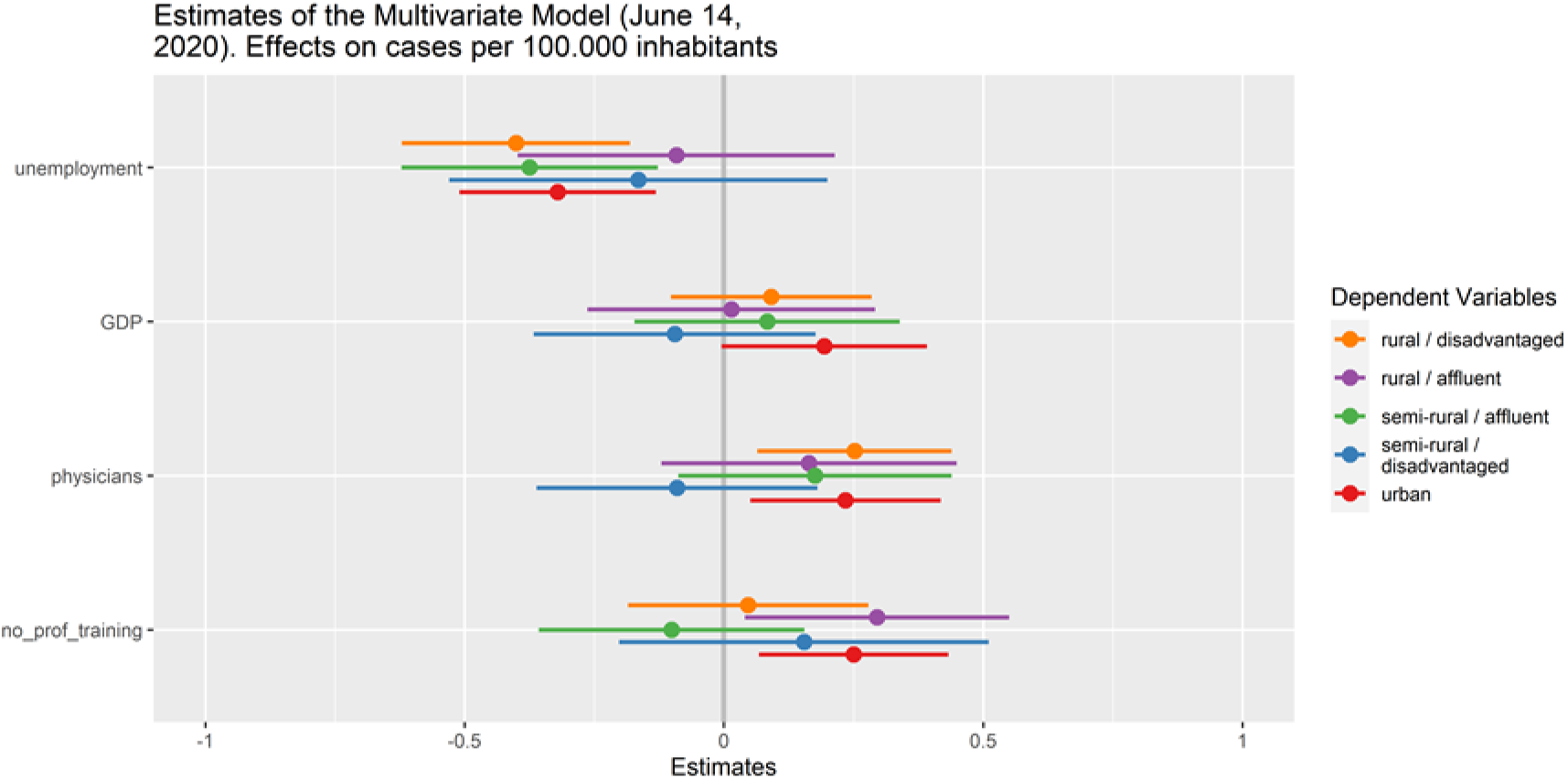
Estimates from different groups (Thünen-type)

The detailed multi-level regression results are shown in table 5: Identically as in previous iterations unemployment has a negative, number of physicians a positive effect on covid-19 cases. Again, low rates of professional training are associated with higher case numbers. The difference between richer and poorer rural zones however becomes highly visible in the multilevel regression and the random effects figures (figure 6). Rural and semi-rural zones with high economic success (category 2 and 3) did have much higher case numbers than other rural zones or even urban centers on average as shown in figure 4.

**Table 5:** Multi-level Regression

|  | Model 1<br>Multilevel Model<br>June 14, 2020 | Model 2<br>Multilevel Model<br>July 7, 2020 |
| --- | --- | --- |
| (Intercept) | 24.727<br>(75.987) | 35.972<br>(78.002) |
| unemployment | -20.810***<br>(2.862) | -19.877***<br>(2.982) |
| GDP | 0.688<br>(0.537) | 0.877<br>(0.558) |
| physicians | 4.025***<br>(0.979) | 3.651***<br>(1.003) |
| no_prof_training | 6.289*<br>(2.711) | 7.118*<br>(2.788) |
| AIC | 5115.763 | 5134.161 |
| BIC | 5143.703 | 5162.101 |
| Log Likelihood | -2550.882 | -2560.080 |
| Num. obs. | 400 | 400 |
| Num. groups: district_type | 5 | 5 |
| Var: district_type (Intercept) | 89.068 | 154.954 |
| Var: Residual | 21273.638 | 22258.950 |

\*\*\*p < 0.001; \*\*p < 0.01; \*p < 0.05
Statistical models

**Figure 6:**
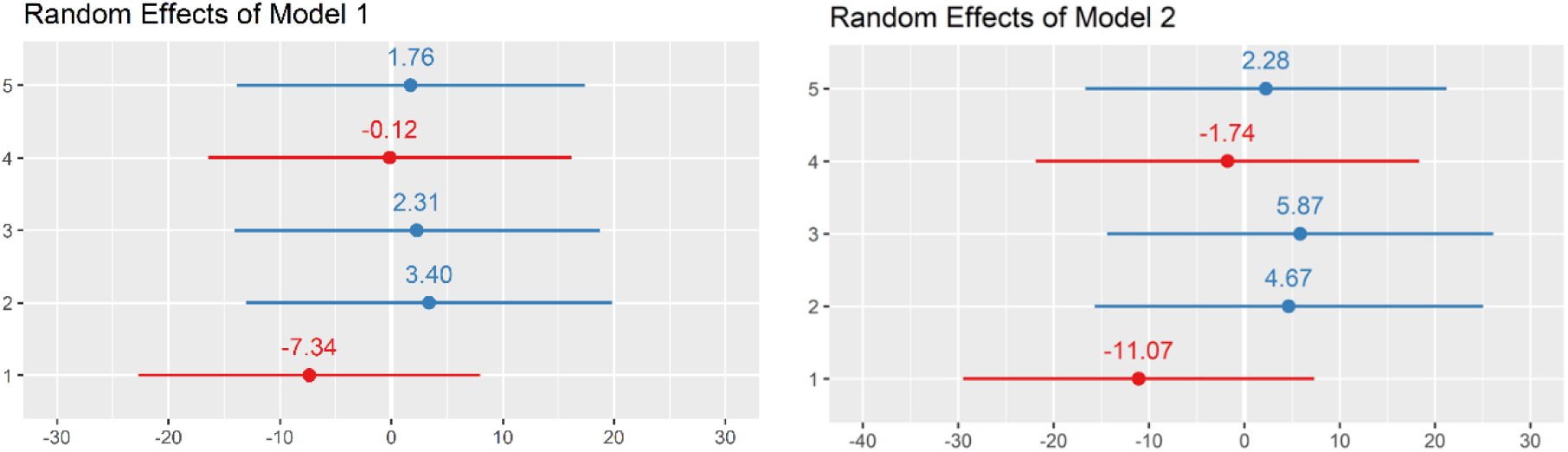
Random effects from model 1 and 2

This pattern of higher case numbers in affluent rural and semi-rural districts did not significantly change between the June and the July measurement, as can be seen in the random effect patterns in figure 6. Affluent and poor rural districts in Germany did experience the pandemic very differently until mid-July with much higher on average infection rates among more affluent districts. Looking at the differences and similarities between the variable significance between all 5 types (figure 5) there is however no clear pattern distinguishing between affluent and disadvantaged districts. The variable “percentage of workers w/o professional training” becomes significant in urban and affluent rural districts. The variable “physicians” indicating the number per capita of general practitioners in the districts becomes significant for urban and disadvantaged rural districts. GDP remains insignificant for all types and unemployment is significant and negatively related for disadvantaged rural, affluent semi-rural and urban districts to case numbers.

While there is a measurable difference between the 5 groups of districts, the ICC difference between groups is also relatively moderate at a 0.7% level.

In almost all models, GDP of districts seems to have little to none effect on the Covid-19 case distribution, except for the second measurement date on July 7 for urban districts. Case numbers are also in general higher were more general physicians are available. But this effect also becomes weaker for urban districts in the second date. This leads to an assumption which should be further tested in the following part of the paper with the data of Berlin: Poverty overall was apparently not an important determinant in the beginning of the outbreak to explain the spreading behavior of Covid-19 in Germany - but is it becoming more important to predict new urban cases in the more recent stage of the outbreak?

Looking at the death rates, the statistical connection between the socio-economic indicators becomes very weak to non-existent. With an adjusted R-squared of below 0.05 and high p-values the included models offer little evidence that mortality in Germany was in anyway meaningfully influenced by or connected to the investigated socio-economic factors on a district level. In the provided models (table 6) unemployment was negatively affecting the death rates for non-rural districts. But due to the models’ weakness, this relationship is highly questionable. Due to the high statistical uncertainty, all selected variables appear to be not systematically related to the mortality rate.

**Table 6.**

| Socioeconomic effects on the death rate |  |  |  |  |  |  |
| --- | --- | --- | --- | --- | --- | --- |
|  | **Model 1**<br>**All Districts**<br>**June 14, 2020** | **Model 2**<br>**Urban Districts**<br>**June 14, 2020** | **Model 3**<br>**Rural Districts**<br>**June 14, 2020** | **Model 4**<br>**All Districts**<br>**July 7, 2020** | **Model 5**<br>**Urban Districts**<br>**July 7, 2020** | **Model 6**<br>**Rural Districts**<br>**July 7, 2020** |
| (Intercept) | 3.197*<br>(1.393) | 3.741<br>(2.946) | 2.914<br>(1.650) | 3.059*<br>(1.363) | 3.369<br>(2.809) | 2.787<br>(1.623) |
| unemployment | -0.165**<br>(0.051) | -0.093<br>(0.098) | -0.196**<br>(0.071) | -0.176***<br>(0.050) | -0.109<br>(0.094) | -0.201**<br>(0.070) |
| GDP | 0.014<br>(0.010) | 0.029*<br>(0.014) | 0.018<br>(0.018) | 0.012<br>(0.009) | 0.027*<br>(0.013) | 0.015<br>(0.018) |
| physicians | 0.029<br>(0.018) | -0.025<br>(0.036) | 0.042<br>(0.021) | 0.033<br>(0.018) | -0.016<br>(0.034) | 0.045*<br>(0.021) |
| no_prof_training | -0.003<br>(0.049) | 0.108<br>(0.100) | -0.039<br>(0.067) | -0.006<br>(0.048) | 0.105<br>(0.096) | -0.038<br>(0.066) |
| R<sup>2</sup> | 0.050 | 0.086 | 0.054 | 0.055 | 0.090 | 0.057 |
| Adj. R<sup>2</sup> | 0.040 | 0.046 | 0.041 | 0.046 | 0.049 | 0.045 |
| Num. obs. | 400 | 95 | 305 | 400 | 95 | 305 |

\*\*\* p < 0.001; \*\* p < 0.01; \* p < 0.05

The fact that mortality rate of Covid-19 in Germany seems not to be meaningfully influenced by poverty indicators is however astounding, given the results of previously conducted national studies in the UK and US at least a small effect was plausible and expected. As this is a remarkable difference to previous Anglo-American studies, these results will be further tested and elaborated on in the second part of the study, looking at the socio-economic data profile of Berlin.

## Evidence from Berlin

Looking at a connected metropolitan area like Berlin in details gives us an opportunity to compare the effects of poverty on Covid-19 in cities and urban areas with the contrasting results from rural zones provided in the previous chapter.

In a series of bivariate examinations shown in table 7, we see that unemployment (SGB II), the number of social assistance recipients (SGB II and XII) as well as population density all remain insignificant until June 14, 2020 for Berlin. On the second measurement date, July 7, 2020 however, we see that all 3 variables shifted further in the direction of statistical significance, making social assistance significant on the 5% level and considerably increasing the determination coefficient of the model.

**Table 7.**

| Socioeconomic effects on cases per 100.000 inhabitants |  |  |  |  |  |  |
| --- | --- | --- | --- | --- | --- | --- |
|  | Berlin Model 1<br>Unemployment<br>June 14, 2020 | Berlin Model 2<br>Unemployment<br>July 7, 2020 | Berlin Model 3<br>Social Assistance<br>June 14, 2020 | Berlin Model 4<br>Social Assistance<br>July 7, 2020 | Berlin Model 5<br>Population Density<br>June 14, 2020 | Berlin Model 6<br>Population Density<br>July 7, 2020 |
| (Intercept) | 144.484* | 122.990 | 136.495** | 124.000* | 154.629*** | 172.818*** |
|  | (53.244) | (60.053) | (41.642) | (45.434) | (23.473) | (27.795) |
| unemployed | 10.926 | 23.649 |  |  |  |  |
|  | (12.363) | (13.944) |  |  |  |  |
| soc.assistance |  |  | 4.223 | 7.696* |  |  |
|  |  |  | (3.126) | (3.410) |  |  |
| Pop.density |  |  |  |  | 0.006 | 0.009 |
|  |  |  |  |  | (0.004) | (0.004) |
| R<sup>2</sup> | 0.072 | 0.223 | 0.154 | 0.337 | 0.237 | 0.296 |
| Adj. R<sup>2</sup> | -0.020 | 0.146 | 0.070 | 0.271 | 0.161 | 0.226 |
| Num. obs. | 12 | 12 | 12 | 12 | 12 | 12 |

\*\*\*p < 0.001; \*\*p < 0.01; \*p < 0.05. Bivariate Regression Models due to limited number of cases

The rate of social assistance reception in Berlin is with an over 95% probability not a random fluctuation in the data and genuinely connected to the observed case occurrence of Covid-19. Also, it should be noted that all 3 indicators are moving simultaneously in the same direction (figure 7) and provide better R-squared values in all later models, indicating a genuine trend in the data, not a plain measurement error caused by fluctuations.

**Figure 7.**
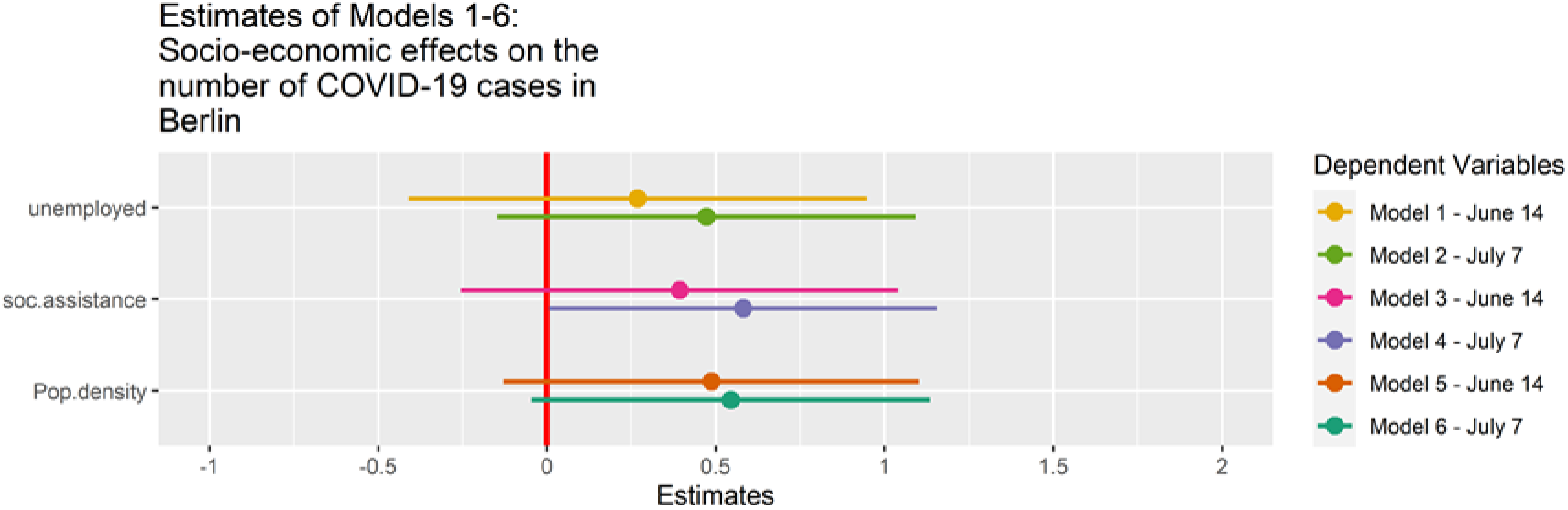

Again, unemployment remains insignificant, but social assistance benefits, which often can be accessed while simultaneously being employed in low qualification jobs becomes significant on the second date. This further hints at the previous observation that the rate of precarious employment – but not unemployment - was connected to higher case numbers in Germany. Considering mortality rates on the other hand, there is again, similar to the data of all German districts, little evidence for a more severe situation within poorer districts of Berlin as demonstrated in table 8. All results on both dates remain statistically insignificant and the overall model parameters are weak. Mortality in Berlin seems not to be systematically affected by the included and considered socio-economic indicators. But since all results are negative, one would assume that whatever indirect effect might exist, it would probably be slightly negative (figure 8). Poverty related indicators thus even hint at potentially lower mortality rates. This could be explained by two aspects: The German healthcare system is due to its universal coverage probably not causing any significant access limitations or different medical procedural outcomes for the average poorer citizen living in deprived districts. Furthermore, because these districts are on average younger than the more affluent districts, they might have an age-related demographic advantage in terms of mortality which is more than compensating the poverty related risks. Covid-19 severity is highly dependent on patient age, so this relation appears plausible (Compare decision tree severity analysis for over 4.000 NYC patients in Petrilli et al. 2020).

**Table 8.**

| Socioeconomic effects on the death rate |  |  |  |  |  |  |
| --- | --- | --- | --- | --- | --- | --- |
|  | **Berlin Model 1**<br>**Unemployment**<br>**June 14, 2020** | **Berlin Model 2**<br>**Unemployment**<br>**July 7, 2020** | **Berlin Model 3**<br>**Social Assistance**<br>**June 14, 2020** | **Berlin Model 4**<br>**Social Assistance**<br>**July 7, 2020** | **Berlin Model 5**<br>**Population Density**<br>**June 14, 2020** | **Berlin Model 6**<br>**Population Density**<br>**July 7, 2020** |
| (Intercept) | 3.345*<br>(1.365) | 3.335*<br>(1.248) | 3.616**<br>(1.096) | 3.471**<br>(0.996) | 3.629***<br>(0.600) | 3.190***<br>(0.567) |
| unemployed | -0.123<br>(0.317) | -0.199<br>(0.290) |  |  |  |  |
| soc.assistance |  |  | -0.062<br>(0.082) | -0.076<br>(0.075) |  |  |
| Pop.density |  |  |  |  | -0.000<br>(0.000) | -0.000<br>(0.000) |
| R<sup>2</sup> | 0.015 | 0.045 | 0.053 | 0.094 | 0.194 | 0.167 |
| Adj. R<sup>2</sup> | -0.084 | -0.050 | -0.041 | 0.004 | 0.113 | 0.084 |
| Num. obs. | 12 | 12 | 12 | 12 | 12 | 12 |

\*\*\* p < 0.001; \*\* p < 0.01; \* p < 0.05. Bivariate Regression Models due to limited number of cases

**Figure 8.**
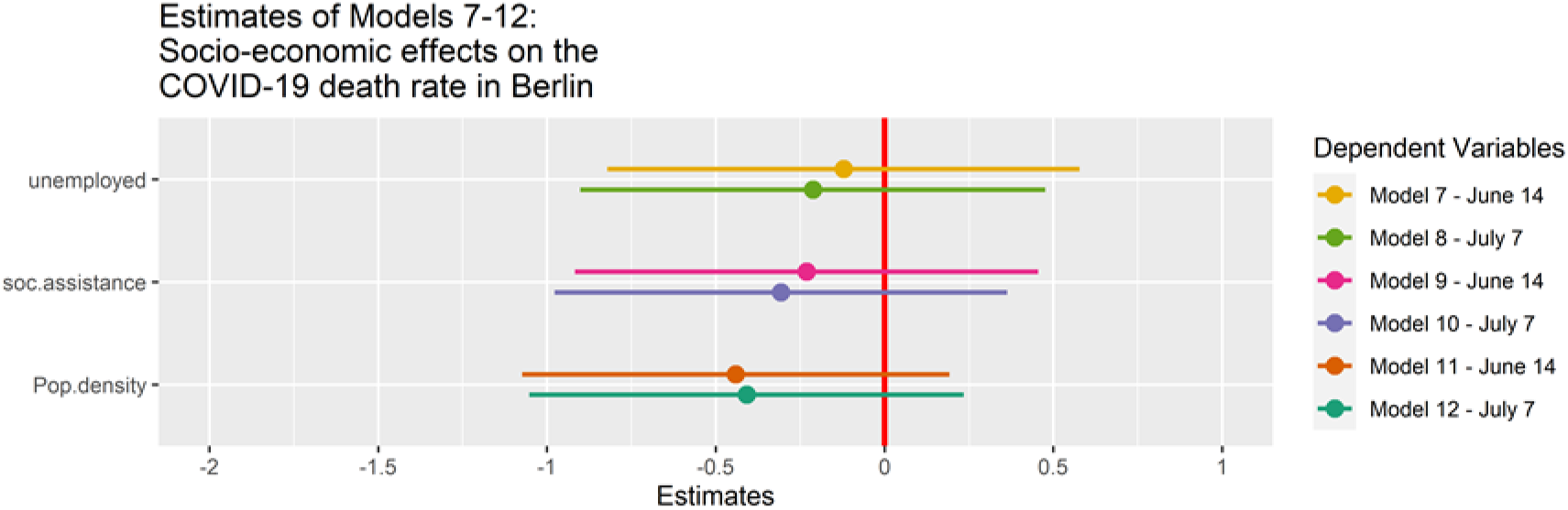

This specific interpretation of the results however needs further proof and a thorough investigation also on a non-aggregated level. Therefore, I caution the usage of this claims without further detailed investigation conducted in separate studies.

## Conclusion

It has been suspected in media and anecdotal commentary that especially in the beginning of the outbreak, young, mobile, and potentially more affluent citizens from the southern parts of Germany were driving the first surge of Covid-19 cases in the country. Returning skiers from holiday sites in Italy played an important role in the early parts of the outbreak. The available socio-economic data the applied regression models based on regional dummies from German districts seem to confirm this connection.

In Germany, the data indicates that the relationship between poverty and cases, but especially the link between poverty and mortality, are much weaker than in the previous Anglo-American studies. There is tentative evidence for a slight increase of influence over time for poverty on case numbers in general, the longer the Pandemic continues. This is highly interesting, as it also directly contradicts the findings of research results from US Districts were poverty became progressively less important for newer cases. Finch and Hernandez Finch (2020) encountered for the US a strong effect of poverty on case numbers in the beginning of the outbreak that diminished over time. The fact that the pattern of poverty-case relationship over time is reversed in Germany indicates that countries might have very distinct and unique interaction patterns for socio-economic variables and infection related data depending on a multitude of socio-economic and political variables.

Another observation for Germany is that the states covering the territory of the former Eastern Democratic Republic, a region with still higher structural unemployment and lower physician numbers and GDP, were affected less by the covid-19 outbreak until June 2020. But the main geographical difference in case distribution was along the north-south divide with highly elevated case numbers in the more affluent southern states. This fact presumably contributed to the negative statistical effect of unemployment on Covid-19 cases in Germany. It also indicates that mobility as principal driving factor of the outbreak was apparently decisive during the early stages of the outbreak.

Despite this overlying trend there are some important hints in the data that correspond to socio-economic effects related to the number of cases: Low qualification work, measured by the proportion of workers without professional training and often associated with manual labor and low quality or even exploitive service jobs, becomes a significant variable in influencing the number of infections in an urban context. In rural districts, there is only in relatively affluent districts a similar effect to be found, showing that the influence of socio-economic factors on the pandemic is highly dependent on the rural-urban structure of each individual district.

Because Germany implemented quick and relatively successful countermeasures against the pandemic and rapidly reduced new case numbers before community transmission became too dominant, these patterns remain prevalent in the distribution of cases on a district level for accumulated data until July 2020. Unlike in other countries, poor districts did neither become excessively hit by the outbreak nor developed higher than average fatality rates. Also, the universal health-care system presumably contributed to the absence of significantly increased mortality rates in poorer districts, in Berlin as well as in Germany as a whole.

Looking at the 5 categories including a systematic separation between affluent and marginalized rural districts, we can see the following pattern: poor rural communities suffered lower case numbers from the outbreak on average, compared to affluent rural, semi-rural and urban districts. Again, this seems to be related to the differences in mobility and connectiveness of these districts.

For Covid-19 cases in Berlin, socio-economic measurements like unemployment and social assistance remain insignificant in the first measurement date but increase their statistical effect on the second date, indicating a potentially increasing effect of socio-economic indicators, the longer the crisis continues. Social assistance became significant in the second measurement date, supporting the evidence from German districts, that especially in the urban context the working poor were more susceptible to higher case numbers, but not the unemployed.

The current non-effect of poverty related indicators on the mortality rates on the other hand seems surprising and in contradiction to previous studies (Finch and Hernandez Finch 2020, Caul 2020). Germany’s and Berlin’s mortality pattern present a very different story compared to the previously mentioned studies from the United Kingdom and the US. A relatively universal and accessible health care system, with less income dependent clinical outcomes and high accessibility for all social groups might be causing this difference. But also, differences in the severity of socio-economic deprivation as well as the very rapid and effective response in reducing case numbers in Germany might have been responsible for the much lower death toll on deprived communities and zones. To evaluate these differences further, I strongly recommend more investigation into this phenomenon in coming studies and more cross-national examinations to study these differences.

## Data Availability

All data is available on request and will be made accessible in a repository prior to submission to a journal

## Footnotes

1 The 63 more affluent rural districts (determined by the Thünen-type) also have a significant effect of professional training on case numbers, as later shown in the multilevel regression (table 4 and figure 5). This effect is however obscured if all rural districts are collectively taken into account. The affluency of rural districts might be connected to having more industrialized farming facilities and more precarious employment often depending on immigrant workers in a specific region. This effect should be tested in more detailed follow-up studies.

## Literature

Blumenthal, D., Fowler, E.J., Abrams, M., Collins, S.R., 2020. Covid-19 — Implications for the Health Care System. The New England Journal of Medicine. https://doi.org/DOI:10.1056/NEJMsb2021088

Caul, S., 2020. Deaths involving COVID-19 by local area and socioeconomic deprivation: deaths occurring between 1 March and 31 May 2020. Office for National Statistics, United Kingdom.

Cooke, T.J., 1999. Geographic context and concentrated urban poverty within the United States. Urban Geography 20, 552–566.

Finch, W.H., Hernandez Finch, M., 2020. Poverty and Covid-19: Rates of Incidence and Deaths in the United States During the First 10 Weeks of the Pandemic. Frontiers in Sociology 5. https://doi.org/doi:10.3389/fsoc.2020.00047

Gaglioti, A., Douglas, M., Li, C., Baltrus, P., Blount, M., Mack, D., 2020. County-Level Proportion of Non-Hispanic Black Population is Associated with Increased County Confirmed COVID-19 Case Rates After Accounting for Poverty, Insurance Status, and Population Density. White Paper - Morehouse School of Medicine.

ILO, 2020. COVID-19 and the world of work: Impact and policy responses (No. Monitor 1st Edition). International Labour Organization.

Kalogeropoulos, A., Suiter, J., Udris, L., Eisenegger, M., 2019. News Media Trust and News Consumption: Factors Related to Trust in News in 35 Countries. International Journal of Communication 13, 3672–3693.

Kenway, P., Holden, J., 2020. Accounting for the Variation in the Confirmed Covid-19 Caseload across England: An analysis of the role of multi-generation households, London and time. New Policy Institute, London.

Levine, J.A., 2011. Poverty and Obesity in the U.S. Diabetes 60, 2667–2668. https://doi.org/10.2337/db11-1118

McFarlane, C., 2020. The urban poor have been hit hard by coronavirus. We must ask who cities are designed to serve. The Conversation.

Mensink, G., Schienkiewitz, A., Haftenberger, M., Lampert, T., Ziese, T., Scheidt-Nave, C., 2013. Overweight and Obesity in Germany. Results of the German Health Interview and Examination Survey for Adults (DEGS1). Bundesgesundheitsblatt 56, 786–794. https://doi.org/10.1007/s00103-012-1656-3

Petrilli, C.M., Jones, S.A., Yang, J., Rajagopalan, H., O’Donnell, L., Chernyak, Y., Tobin, K.A., Cerfolio, R., Francois, F., Horwitz, L.I., n.d. Factors associated with hospitalization and critical illness among 4,103 patients with Covid-19 disease in New York City. medRxiv. https://doi.org/10.1101/2020.04.08.20057794

Rachele, J.N., Kavanagh, A.M., Badland, H., Giles-Cort, B., Washington, S., Turrell, G., 2015. Associations between individual socioeconomic position, neighbourhood disadvantage and transport mode: baseline results from the HABITAT multilevel study. Journal of Epidemiology and Community Health 69, 1217–1223. https://doi.org/doi:10.1136/jech-2015-205620

Robles, S., Simington, J., Shaefer, H.I., 2019. Index of Deep Disadvantage: Technical Documentation. Retrieved from the University of Michigan Poverty Solutions Initiative. University of Michigan.

